# ChatENT: Augmented Large Language Model for Expert Knowledge Retrieval in Otolaryngology - Head and Neck Surgery

**DOI:** 10.1101/2023.08.18.23294283

**Authors:** Cai Long, Deepak Subburam, Kayle Lowe, André dos Santos, Jessica Zhang, Sang Hwang, Neil Saduka, Yoav Horev, Tao Su, David Cote, Erin Wright

**Affiliations:** University of Alberta; Klavier AI; Alberta Machine Intelligence Institute; Fudan University, China; Sengled AI

## Abstract

**Background:** The recent surge in popularity of Large Language Models (LLMs), such as ChatGPT, has showcased their proficiency in medical examinations and potential contributions to medical education. However, LLMs possess inherent limitations, including inconsistent accuracy, specific prompting requirements, and the risk of generating harmful hallucinations. A domain-specific, fine-tuned model would address these limitations effectively.

**Methods:** OHNS-relevant data was systematically gathered from open-access internet sources and indexed into a database. We leveraged Retrieval-Augmented Language Modeling (RALM) to recall this information and used it for pre-training, which was then integrated into ChatGPT 4·0, creating a OHNS specific knowledge Q&A platform known as ChatENT.

**Findings:** ChatENT showed enhanced performance in the analysis and interpretation of OHNS information, outperforming ChatGPT 4.0 in both the Canadian Royal College OHNS sample examination questions challenge and the US board practice questions challenge, with a 58.4% and 26·0% error reduction, respectively. ChatENT generated fewer hallucinations and demonstrated greater consistency.

**Interpretation:** To the best of our knowledge, ChatENT is the first specialty-specific LLM in the medical field. It appears to have considerable promise in areas such as medical education, patient education, and clinical decision support. The fine-tuned model has demonstrated the capacity to overcome the limitations of existing LLMs, thereby signaling a future of more precise, safe, and user-friendly applications in the realm of OHNS.

**Funding:** The authors received no financial support for the research, authorship, and/or publication of this project.

## Introduction

Large language models (LLMs) are artificial intelligence (AI) systems that are trained on text-based human knowledge, derived from articles, books, and other internet-based content. These advanced artificial intelligence (AI) models effectively grasp language syntax, semantics and patterns, enabling the generation of dynamic, coherent text-based responses.^1–3^ With their advanced information structuring capabilities, LLMs have revolutionized many fields, including medicine—offering applications with significant healthcare impact while helping mitigate medical errors.^4–6^

In medicine, LLMs have been applied to many realms, including medical education, clinical decision support, and analysis of medical images and patient data.^5,7–11^ Furthermore, recent evaluations on LLMs’ effectiveness in generating medical reports and responding to consults showed improvements in administrative efficiency. Specific area of focus include information management in radiology and pathology.^12–15^

Several LLMs have been tailor-made for medical contexts to optimize AI performance in healthcare. Notably, by leveraging Bidirectional Encoder Representations from Transformers (BERT), specialized models like BioBERT and ClinicalBERT have emerged, designed using biomedical texts to enhance language comprehension within the medical sphere.^16^ However, these models sometimes grapple with managing contextual data.

Several approaches have been proposed to address unique healthcare needs. Relevant examples include an expert system for psychiatric consultations and knowledge graphs for neurologic subarachnoid hemorrhage predictions.^17,18^ Singhal et al. introduced Med-PaLM, which achieved good results in knowledge retrieval and clinical decision support tasks, but still fell short compared to human clinicians’ performance.^16^ Med-PaLM was assessed on a human evaluation framework using traits such as factuality, comprehension, reasoning, possible harm and bias^19^. Jeblick et al focused on exploring LLMs’ potential in simplifying intricate medical reports in the context of radiology.^20^ A generative model was developed in UK to analyze Electronic Health Records to make medical risk forecasts.^21^

Though LLMs generally come with policies that restrict their use in high-risk sectors like legal and healthcare services, recent advancements, particularly in models like ChatGPT and Bard, have shown great potential for healthcare applications.^3,22^ However, the inherent risks associated with LLMs such as truthfulness, bias, toxicity, and other societal risks, necessitate further research.^3^

ChatGPT is currently one of the most popular LLMs used and tested specifically among medical specialties. It has been evaluated broadly through physician-generated questions and physician-based appraisal across seventeen specialties for accuracy and reliability.^23^ For otolaryngology— head and neck surgery (OHNS) in particular, ChatGPT was assessed as an informational resource for OHNS patients for safety, accuracy and comprehensiveness, as well as on the Royal College of Physicians and Surgeons of Canada’s OHNS board exam based on concordance, validity, safety, accuracy, where it showed satisfactory performance.^24,25^ However, given LLMs’ potential biases, misuses and their limitations in producing complete and factually accurate outputs, ethical considerations such as data privacy, accountability, and fairness should be accounted for when working with LLMs in healthcare.^26,27^ Some attempts have been made to address these issues, including developing LLMs that focus on specific areas.^28^ Zakka et al^29^ published a retrieval-augmented oriented language model for generalized clinical medicine utilizing external tools, including search engines.

Generally, building an LLM may take a great amount of resources and time.^30^ There is a growing interest and need for Green AI, where those using and developing AI models are held more accountable for their carbon impacts.^31^

To our knowledge, no medical specialty-specific LLMs, such as those centering on otolaryngology or other fields, have been published. Every specialty possesses its unique knowledge base, with abundance of acronyms and synonyms, necessitating meticulous curation and adaptation. A framework with demonstrated capacity to overcome the limitations of existing LLMs would be crucial to a future of more precise, safe, and user-friendly applications in diverse realms of clinical practice for both clinicians and patients. We hereby propose a cost-efficient, simplified approach to construct a medical specialty knowledge domain AI, demonstrated by ChatENT.

## Methods

Figure 1 shows the flow of operations, from question entry to answer generation. The first step is a one-time preparation of the ChatENT Knowledge Base: we split this corpus of text into one-page-length chunks of text, and feed each chunk to OpenAI’s embedding model (text-embedding-ada-002) to get a vector representation (a list of 1,536 real numbers) of each chunk. This is shown in the bottom left of the diagram. When a question is entered, a vector for it is also computed, and compared against all the knowledge base chunk vectors. The 5 knowledge base chunks with the most similar vectors get selected. The text associated with these, along with the question and instruction (prompt text) to ChatGPT, get assembled into a full query. This query is sent to ChatGPT which then generates the answer.

**Figure 1.**
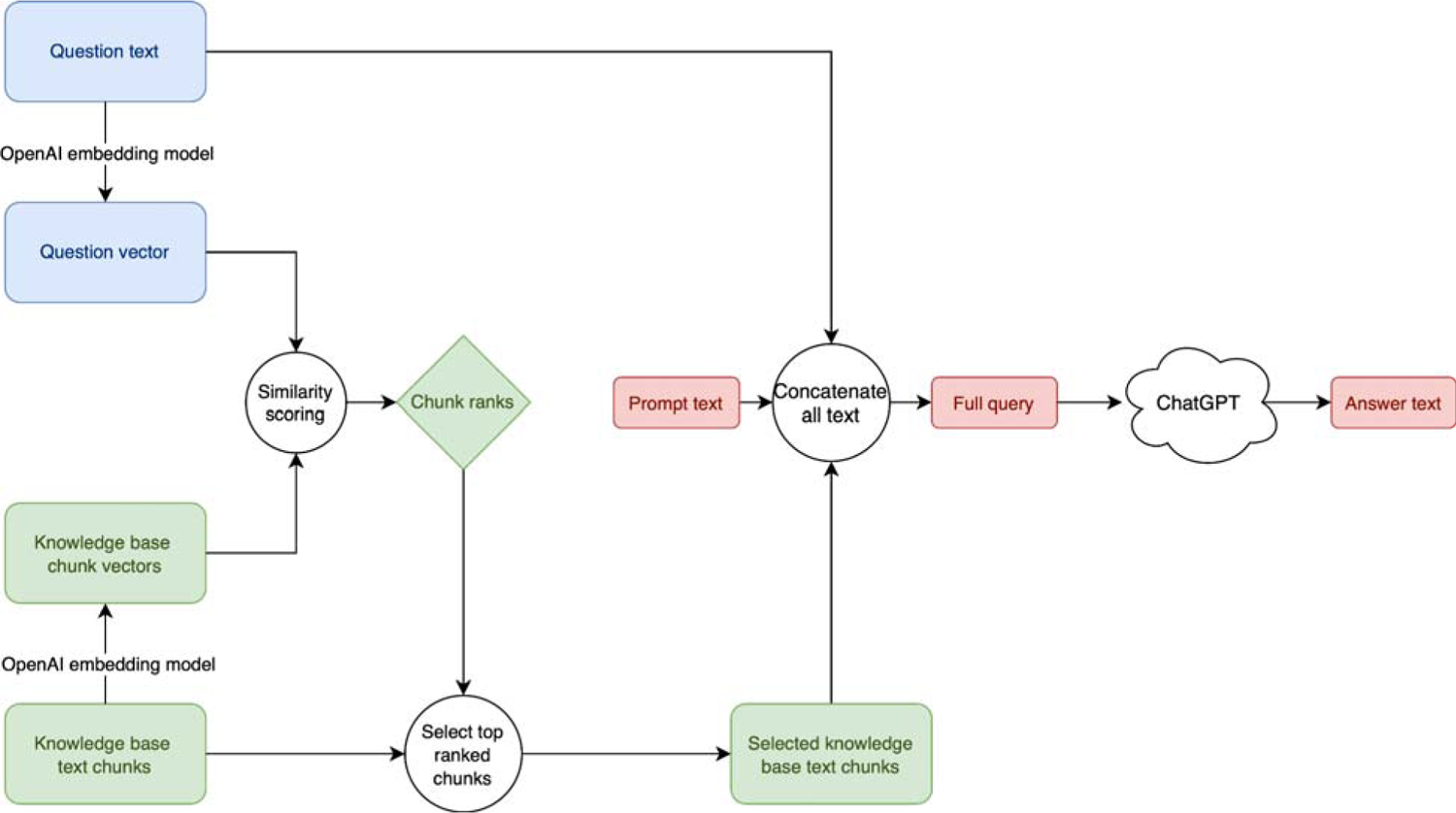
System flow diagram.^a a^Green: Knowledge-relevant steps; Blue: Question-relevant steps; Red: Answer relevant steps.

We improve upon vanilla ChatGPT Q&A by adding a preparatory stage to the process that augments the model with specialized domain knowledge text. In this stage, a separate machine-learning approach is used to retrieve text relevant to the question posed from ChatENT’s Knowledge Base. This retrieved text is pre-pended to the question, and ChatGPT is prompted to answer the question using the information in this preface. We use ChatGPT’s subsequent response as the answer for evaluation in this study.

We do not train ChatGPT on the CEKB, for that is prohibitively expensive even if we could do it, and furthermore is likely not the best approach—see the following section that discusses this.

Neither do we have the option to include the entire CEKB as a preface to the question posed— there is a limit to the size of the full submission to ChatGPT, which ranges from 4,096 tokens (∼10 pages) to 32,768 tokens (∼80 pages) depending on the model version used.

So we pick 6 pages or so of the most relevant content in the CEKB to send ChatGPT, using their 10-page model. This allows enough space for the question itself and ChatGPT’s response (which all need to fit within the 10 pages). These picks are made using the machine-learning technique of *embedding vector similarity search*.

Before employing that technique, we first chunk the CEKB into contiguous blocks of text, each about one page long. This results in more than 20,000 chunks of text. The task is then, for any given question, to pick the 6 or so chunks that are most relevant to that question. Embedding vector similarity search works by representing each of these chunks of text with an embedding vector (a sequence of real numbers). These embedding vectors have the property that text with similar semantic meaning have low cosine distance between their respective embedding vectors.

OpenAI’s text-embedding-ada-002 model ^32,33^ was used to compute the embedding vector for each chunk. When ChatENT receives a user question, that question is also run through the same text-embedding-ada-002 model, giving a single embedding vector representing the question’s semantic meaning. The cosine similarity is then calculated between the question’s embedding vector and the embedding vectors for each chunk in the CEKB. We then pick the 6 or so chunks with the greatest cosine similarity. Hence the name *embedding vector similarity search*.

The picks are then concatenated to create the preface in the full query that is submitted to ChatGPT, as explained at the start of this section.

## Results

The effectiveness of ChatENT was evaluated using two distinct types of queries: open-ended short-answer questions and multiple-choice questions. These formats best mirror the primary types of questions learners encounter in exam settings and emphasize different strengths of LLMs. Success in the former necessitates a comprehensive understanding of the topic and mimics real life clinical use, while the latter demands clinical reasoning and in-depth knowledge.

### Open-ended short answer questions

ChatENT was assessed using sample questions from the Canadian Royal College of Physicians and Surgeons. The CVSA (Concordance, Validity, Safety, and Accuracy) model served as the evaluation metric for the LLM’s performance. The results from this assessment were promising as demonstrated in Figure 2.

**Figure 2.**
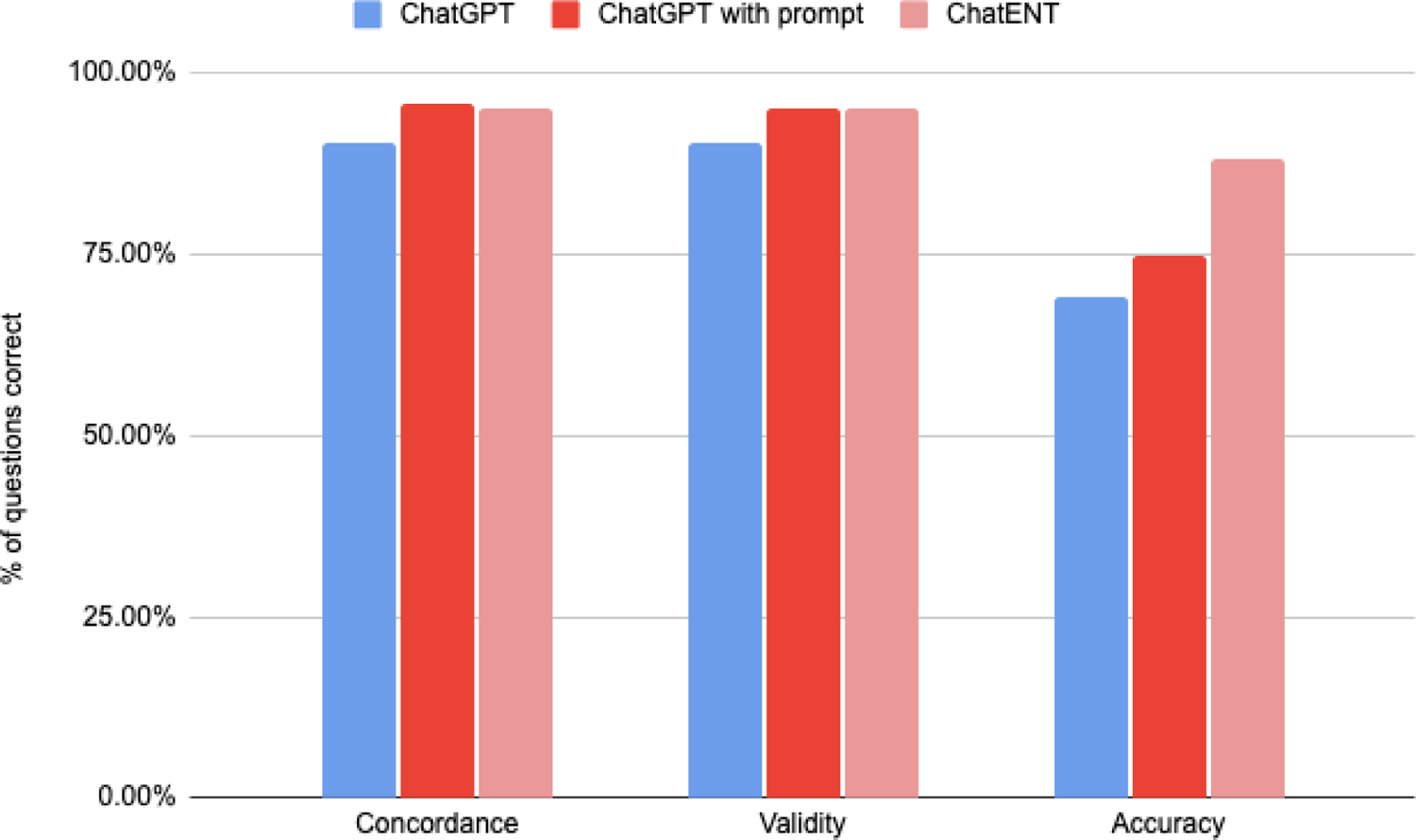
Performance of ChatGPT, ChatGPT with prompting, and ChatENT on questions from a sample Royal College certifying examination for OHNS.

### Concordance

ChatENT demonstrated similar levels of concordance with ChatGPT. Given that the principal dataset used for language training is predicated on a massive dataset of many parameters, it was anticipated that ChatENT’s concordance with ChatGPT would align with expectations based on the characteristics of language-based prediction models.

### Validity

ChatGPT4.0 presented a commendable degree of validity, amounting to 90·5%. However, it is noteworthy that ChatENT surpassed this already high benchmark by achieving an outstanding validity of 95·7%. This suggests that the majority of responses generated by ChatENT is aligned with widely accepted scientific views, grounded in evidence-based, scientifically verified conclusions, or concordant with the consensus among OHNS experts and senior staff.

### Safety

The safety component of this comparative analysis deserves particular attention, Consumer tools like ChatGPT were not trained on the entire corpus of biomedical data, nor was it tested or trained by experts in biomedicine.^34^ In contrast to ChatGPT, which had three potential safety red flags identified in the responses it generated, ChatENT maintained a robust safety record with zero instances of alarming red flags in our testing. This underlines the enhanced safety features of ChatENT, highlighting its reliability when compared to other models.

Sample red flags from ChatGPT and contrasting answers from ChatENT are documented in Table 1.

**Table 1.** Sample question and answers from ChatGPT and ChatENT.

| Question | ChatGPT | ChatENT |
| --- | --- | --- |
| 1. You are performing an osteoplastic flap procedure using a bicoronal approach for drainage of mucoceles.<br>In which soft tissue planes will you elevate the bicoronal flap?<br>Once the frontal bone is exposed, list three techniques to identify margins of the frontal sinus prior to making the bony cuts for the osteoplastic flap. | [...] Three techniques to help with this process are:<br>Digital palpation: One of the simplest methods to identify the margins of the frontal sinus is by using digital palpation. By carefully palpating the frontal bone, you can often feel the transition between the thicker bone of the frontal sinus and the thinner bone of the calvarial vault. The borders of the frontal sinus can be marked based on these tactile findings<br>[...] | Once the frontal bone is exposed, three techniques to identify the margins of the frontal sinus prior to making the bony cuts for the osteoplastic flap include:<br>- Caldwell view X-ray obtained from a distance of 6 feet: This radiographic view helps visualize the frontal sinus and its margins.<br>- Navigation system: Image-guided navigation equipment can be used to accurately identify the position of the frontal sinus and guide the surgeon during the procedure.<br>- Shining an endoscope transnasally: By inserting an endoscope through the nasal cavity, the surgeon can visualize the frontal sinus and its margins directly. |
| 2. Name three techniques used to prevent or cure pharyngoesophageal spasm after total laryngectomy. | [...] Pharyngeal dilation: This procedure involves the insertion of a dilator into the pharynx to gradually stretch the narrowed or spasmodic area, allowing for improved swallowing. This can be performed either through a rigid or a balloon dilator, depending on the patient's | The three techniques used to prevent or cure pharyngoesophageal spasm after total laryngectomy are:<br>1. Cricopharyngeal Myotomy: This procedure involves the surgical cutting of the cricopharyngeus muscle to relieve the spasm. |
|  | specific needs. | 2. Pharyngeal Plexus Neurectomy: Although not widely used, this technique involves the removal of nerve branches entering the lateral wall of the pharynx to relax the constrictor muscles.<br>3. Preservation of Inferior Constrictor: During the laryngectomy, as much of the inferior constrictor muscle (including the cricopharyngeus and thyropharyngeus) is preserved as possible to prevent postoperative spasm. |
| 3. A patient presents with an anterior glottic web. Name five treatment options, used alone or in combination, for the management of this lesion. | Laryngotracheal reconstruction: In severe cases, where the web is causing significant airway obstruction, a laryngotracheal reconstruction may be necessary. This procedure involves surgically enlarging the airway and reconstructing the vocal cords to restore normal function. | [...]5. Laryngotracheal reconstruction (LTR) or cricotracheal resection (CTR): In cases where the web is extensive and associated with subglottic stenosis, a more extensive surgical procedure may be necessary. LTR or CTR involves resecting and reconstructing the affected area to restore a functional airway.[...] |

### Accuracy

A comparative analysis of the accuracy of ChatGPT and ChatENT was conducted, as shown in Figure 2. When used without a predefined prompt, ChatGPT achieved an accuracy score of 69.10%, falling just below the acceptable passing threshold of 70.00%. In contrast, ChatENT displayed a superior performance, reaching a remarkable accuracy score of 87.23%. This constitutes a 58.4% error reduction over the unprompted ChatGPT.

All three models were evaluated by their concordance, validity, and accuracy according to the CVSA model as proposed in a previous study. On average ChatENT performed the best, followed by ChatGPT with prompting and ChatGPT, respectively.

All three models were evaluated for safety issues, according to the CVSA model proposed in a previous study. ChatGPT had the most answers with safety issues, followed by ChatGPT with prompting and none from ChatENT.

### Multiple choice questions (MCQ)

Further, we challenged ChatENT with OHNS multiple choice questions, namely U.S. board practice questions which were widely used by US senior otolaryngology - head and neck surgery residents.

The MCQ question test banks consist of 8 sections, each with 20 questions from subspecialties including general ENT, Head and Neck Surgery, Laryngology, Otology-Neurotology, Facial Plastics Surgery, Pediatrics and Rhinology. The question bank was challenged by ChatGPT, ChatENT respectively. An overall human performance score was calculated using the average percentage of human testor scores correctly on each question, with public available data derived from the question bank.^35^ The test participants are generally senior year OHNS residents who are preparing for the ABOHNS board examination. The overview of the results is summarized in Figure 3.

**Figure 3.**
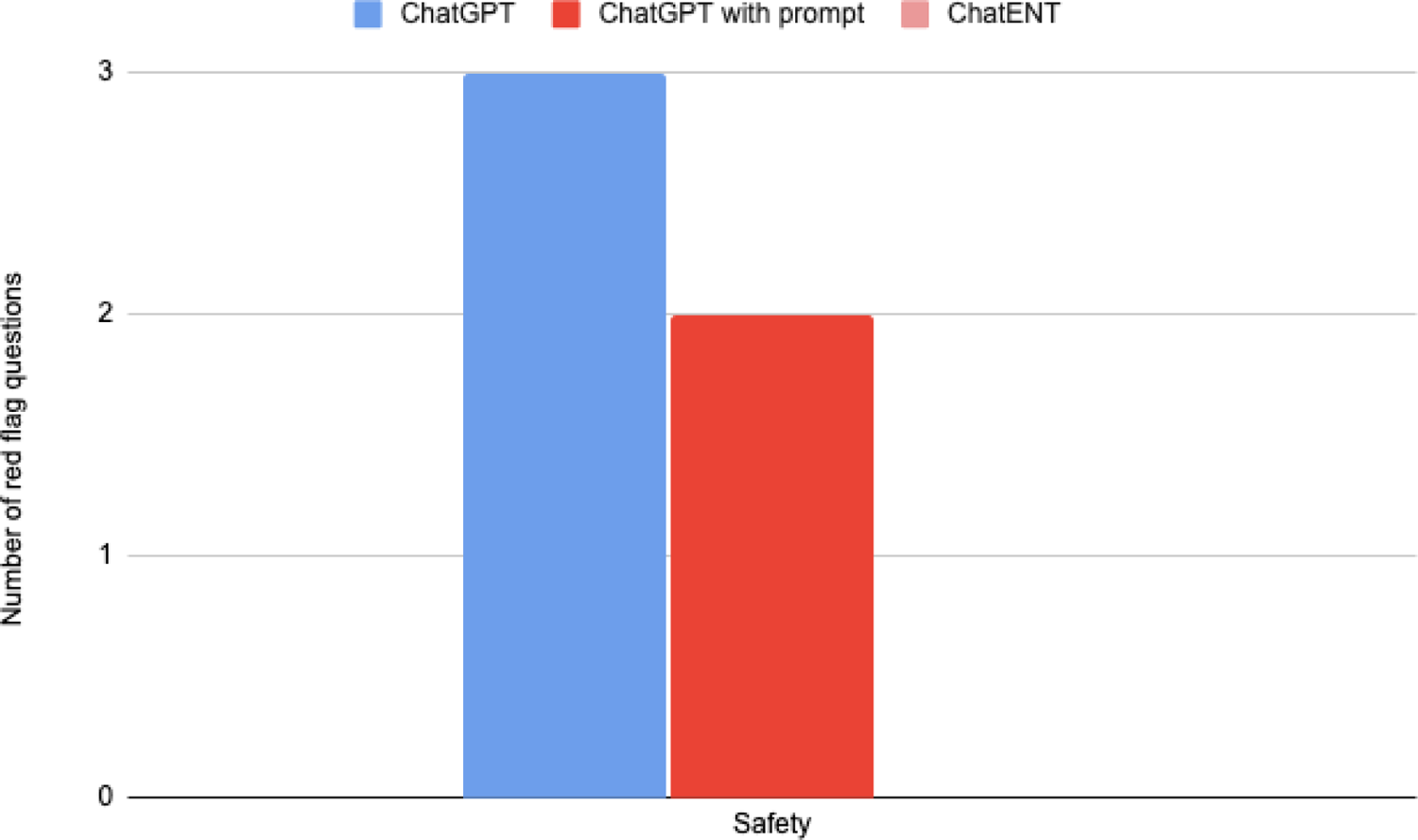
Performance of ChatGPT, ChatGPT with prompting, and ChatENT on questions from a sample Royal College certifying examination for OHNS.

When evaluated against sample OHNS multiple-choice board questions, ChatENT performed better than ChatGPT across all specialties except for Basic Science and Pediatrics, and equally for Head and Neck Surgery. Figure 4 displays results according to worsening ChatENT performance from left to right. Notably, ChatENT’s highest scores were in otology-neurotology (90% vs 75%) and lowest in pediatrics (70% vs 75%). Overall, ChatENT scored better than ChatGPT with 80% correct compared to ChatGPT’s 73%, representing a 26% error reduction.

**Figure 4.**
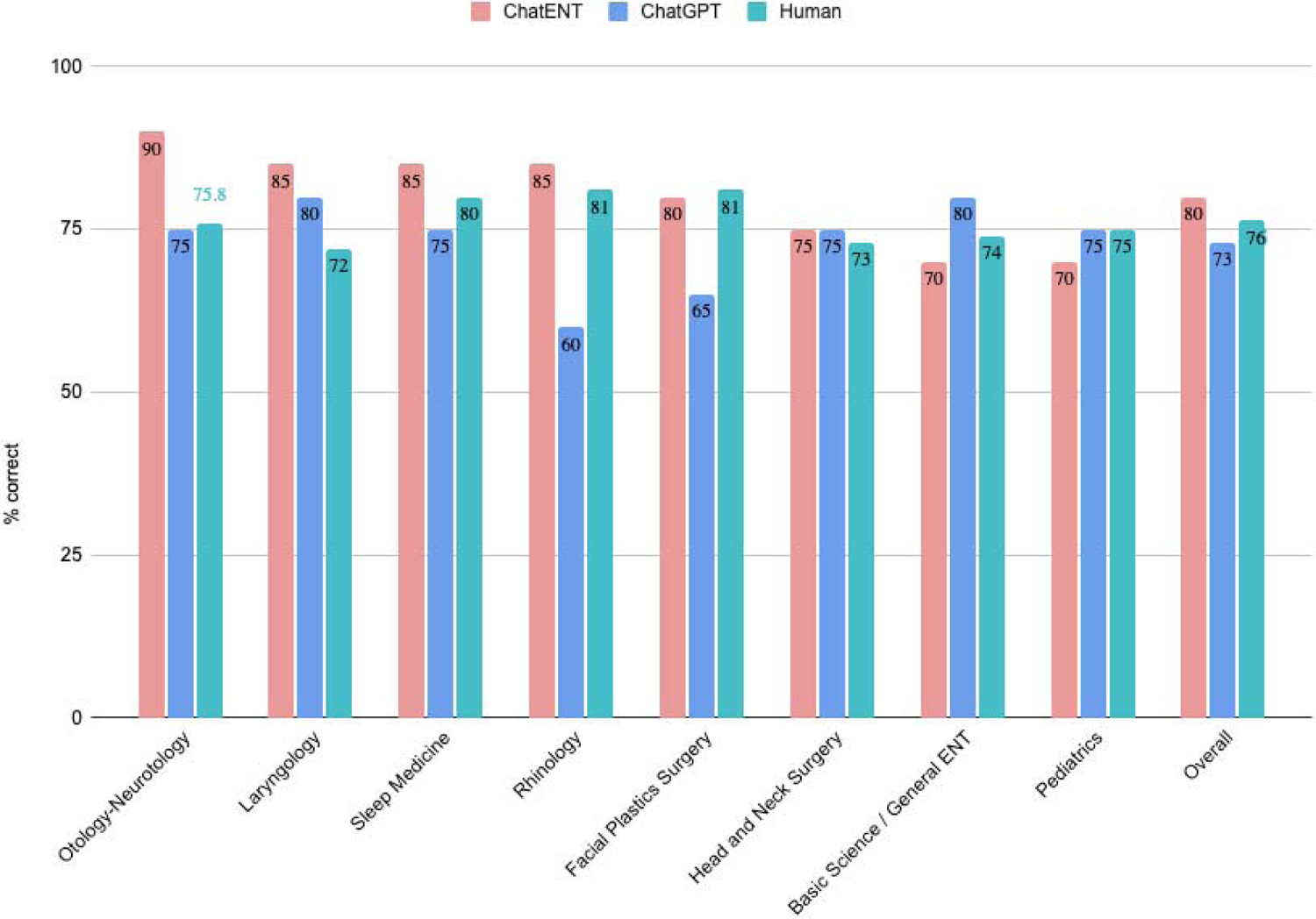
Performance of ChatENT and ChatGPT on subspecialty questions from American Board of Otolaryngology Head and Neck Surgery (ABOHNS) examination practicing question bank.

**Figure 5:**
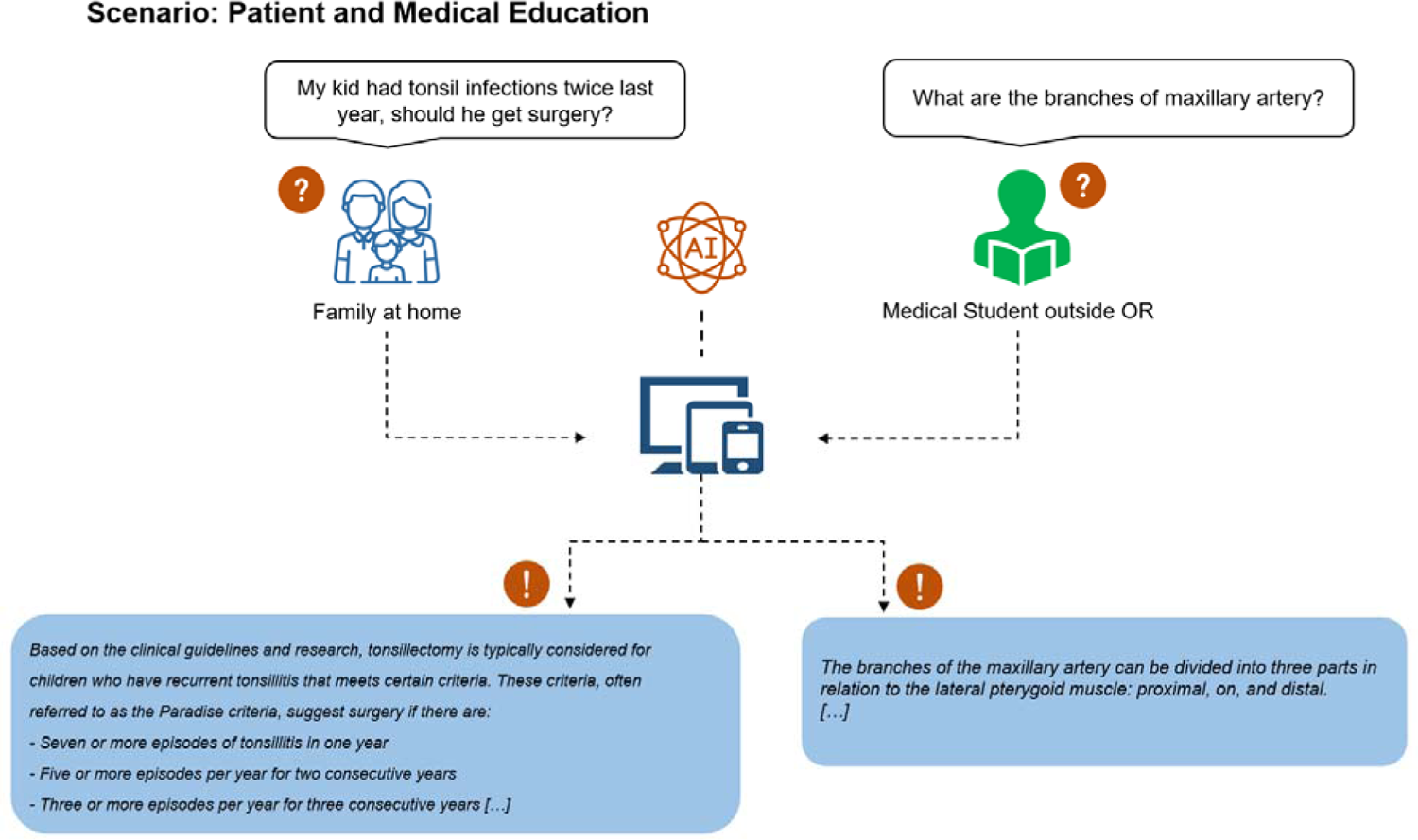
Sample illustration of ChatENT utilization in patient and medical education.

**Figure 6.**
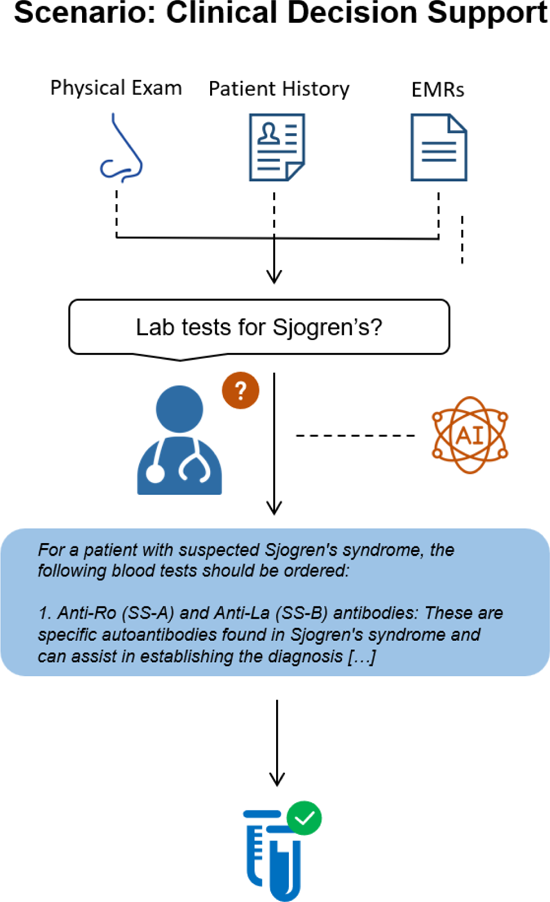
Sample illustration of ChatENT utilization in future clinical decision support.

Additionally, ChatENT appears to perform better than the average human. Notably, the sections where the greatest differences are seen is in otology-neurotology, laryngology, and sleep medicine, where the difference in score is greater than 5% in these three categories. Overall, ChatENT scored better with 80% compared to a human-based score of 76%, representing an error reduction of 17%.

Overall, ChatENT demonstrated superior performance than both ChatGPT and human testors.

## Discussion

ChatENT is a pioneering entity in the realm of artificial intelligence models, being the first to specialize in domain-specific knowledge in a surgical specialty. It exhibited superior performance to ChatGPT4.0 when tested on both multiple-choice and short answer questions sourced from board examination materials.

Upon evaluation with short-answer questions (as illustrated in the attached sample questions), the performance of the system exhibited comparable concordance and validity. This consistent performance can be attributed to the robustness of the overarching LLM framework. However, CHatENT demonstrated enhanced accuracy with significant reduction of safety red flags and errors. Of particular note was a marked reduction in safety-related issues, which are of paramount concern in healthcare applications. This highlights the potential advantages of domain-specific LLMs in shaping optimized scenarios tailored for the healthcare sector.

Further, when subjected to multiple-choice questions (MCQs), the system yielded satisfactory outcomes, outperforming ChatGPT with a commendable 26% reduction in errors. Yet, it is important to note that both LLMs didn’t significantly surpass the performance of human candidates. A potential reason for this is that MCQs frequently contain confounding or superfluous information that can potentially misguide the LLM. When these questions underwent a refining process for optimized comprehension, the retrieved information didn’t resonate as coherently as it did with the short-answer queries. However, it is noteworthy that the system marginally outperformed human testers. This highlights the system’s potential applicability across a range of healthcare domains.

Our platform offers three distinct advantages over the straightforward one-step querying of an LLM:

### Up-to-date Knowledge Base

Our specialized knowledge base is both more expansive and current compared to the LLM’s training data. Even if LLM training dataset compilers diligently incorporate all pertinent sources from our field, they are limited by their publication cut-off, which is September 2021 for GPT-4. With our system, newly minted best-practice articles can be seamlessly integrated into our knowledge base without necessitating any modifications to the LLM itself.

### Quality Assurance

The innate clinical understanding of an LLM can be inconsistent. The sheer volume of text required for Large Language Models might encompass outdated or discredited treatment methodologies and suggestions. Even if an LLM is fine-tuned using recent content, like our knowledge base, it remains vulnerable to lapses. Our model draws focus on the prescreened knowledge base to provide evidence based answers, which is evidenced by the decrease of redflags in our test.

### User-Centered Styling

Supplying the LLM with text extracted from the knowledge base optimally primes it for appropriate responses. Specifically, our strategy yields answers that not only encapsulate the required technical minutiae, but also employ a clinical writing style that lucidly and succinctly conveys the information. We can adapt different styles for different end users to meet their unique needs, for example, for clinical users, a concise answer with only relevant information is preferred, for patients, a more detailed answer in lay language will be more beneficial.

This study underscores the potential applications of specialty-specific artificial intelligence models, including medical education, patient education and decision support (summarized in Table 2).

**Table 2:** Summary of identified advantages, example questions, and example answers pertaining to each advantage.

|  | Medical education | Patient education | Decision support |
| --- | --- | --- | --- |
| Example question | What are the segments of the facial nerve? | My kid had a tonsil infection twice last year, should he get surgery? | What type of lab test should I order for a patient with suspected Sjogren's? |
| Example answer | *The facial nerve is conventionally divided into intracranial (cisternal), intratemporal, and extratemporal portions.*<br>*1. Intracranial (Cisternal) Portion: This segment is approximately 24 mm long and courses anterolaterally through the cerebellopontine angle to the porus of the internal auditory meatus (IAM).*<br>*2. Intratemporal Portion: This segment is 28-30 mm long and is subdivided into meatal, labyrinthine, tympanic (horizontal), and mastoid (vertical) segments.*<br>*- Meatal Segment: Runs in the IAM.*<br>*- Labyrinthine Segment: Runs laterally to the geniculum and the geniculate* | *Based on the clinical guidelines and research, tonsillectomy is typically considered for children who have recurrent tonsillitis that meets certain criteria. These criteria, often referred to as the Paradise criteria, suggest surgery if there are:*<br>*- Seven or more episodes of tonsillitis in one year*<br>*- Five or more episodes per year for two consecutive years*<br>*- Three or more episodes per year for three consecutive years*<br>*Given that your child has had two episodes of tonsillitis last year, it does not meet the criteria for surgery based on frequency. However, the decision for surgery should also take into account the severity of each* | *A: For a patient with suspected Sjogren's syndrome, the following blood tests should be ordered:*<br>*1. Anti-Ro (SS-A) and Anti-La (SS-B) antibodies: These are specific autoantibodies found in Sjogren's syndrome and can assist in establishing the diagnosis.*<br>*2. Rheumatoid Factor (RF): This is often elevated in patients with Sjogren's syndrome.*<br>*3. Antinuclear Antibodies (ANA): These are often present in autoimmune conditions, including Sjogren's syndrome.*<br>*4. Immunoglobulin levels: Specifically, a low level of IgM may indicate an increased risk of progression to* |
|  | ganglion.<br>- *Tympanic (Horizontal) Segment: Runs backwards from the geniculate ganglion to the second genu.*<br>- *Mastoid (Vertical) Segment: Runs from the second genu to the stylomastoid foramen.*<br>3. *Extratemporal Portion: This segment runs forwards into the parotid gland where it usually divides into temporofacial and cervicofacial branches.* | *episode, how well infections have responded to medical therapy, and quality of life issues such as missed school days.*<br>*It's important to have a discussion with your child's healthcare provider, who can consider all these factors and provide a personalized recommendation.* | *malignancy in Sjogren's syndrome.*<br>*Remember, these tests are part of the diagnostic process and should be interpreted in conjunction with the patient's symptoms, clinical history, and other diagnostic procedures such as minor salivary gland biopsy, Schirmer's test, and imaging studies.* |

### Patient Education

A significant advantage of ChatENT lies in its ability to streamline the educational process. It can efficiently provide accurate information, eliminating the need to sift through guidelines, textbooks, or academic papers. This feature is particularly useful in fields such as aesthetic surgery, where patients might feel reluctant to pose questions to another human being.

### Medical Education

A patient-oriented version of ChatENT can be developed to facilitate easy access to health information in plain language. It can eliminate red flags potentially harmful to patients, thereby improving patient understanding and safety.

### Clinical Decision Support

ChatENT can function as a valuable resource for clinicians, particularly in rural and community settings where they might encounter unfamiliar cases. It serves as a readily accessible, reliable reminder, thus enhancing the decision-making process.

### Limitations

We acknowledge several limitations in our study. Firstly, our testing objects for the short-answer question bank were limited, which might not have provided a comprehensive evaluation.

Secondly, the knowledge database, being a dynamic entity, is continually undergoing improvements, its current state might not be its most refined or comprehensive version. Additionally, the population of individuals making up the human-derived score we utilized for subspecialty analysis was from. Thus, the human-derived score may not appropriately reflect a population that is attempting to score to the best of their ability. Finally, we did not extensively assess the patient-centered user interface due to time limits, a vital component for user experience and interaction. This omission might have prevented us from capturing the nuances of how end-users, especially patients, would engage with and experience the system. Future research and iterations of our study should aim to address these shortcomings to enhance the robustness and applicability of our findings.

## Conclusion

We propose a cost-effective method to construct a domain-specific AI for medical specialties, as demonstrated with our ChatENT using augmented retrieval. ChatENT exhibited superior performance on ENT exam questions, excelling in both open-ended short-answer questions and multiple-choice questions. Moreover, it displayed significantly fewer hallucinations. This tool has potential applications in various facets of clinical practice, including medical education, decision support, and patient education. Future endeavors will focus on integrating other AI models into the LLM to broaden its capabilities, for example, incorporating computer vision for medical imaging interpretation and automatic speech recognition for clinical conversation analysis in real time.

## Data Availability

All data produced in the present study are available upon reasonable request to the authors

## Notes

### Competing Interest Statement

The authors have declared no competing interest.

### Funding Statement

This study did not receive any funding

### Summary of Updates

Figure and table revised.

